# Community Assessment of School-Based Mass Drug Administration Program for Soil-Transmitted Helminths and Schistosomiasis in Nigeria

**DOI:** 10.1101/2023.03.06.23286829

**Authors:** Folahanmi T. Akinsolu, Olunike Abodunrin, Mobolaji Olagunju, Ifeoluwa E. Adewole, Nurudeen Rahman, Anita Dabar, Diana W. Njuguna, Islamiat Y. Shoneye, Abideen Salako, Oliver C. Ezechi, Orsolya Varga, Olaoluwa P. Akinwale

## Abstract

**Background:** Neglected tropical diseases, such as soil-transmitted helminths and schistosomiasis, are prevalent in sub-Saharan Africa, particularly in Nigeria. Mass drug administration is the primary control intervention, but the coverage and utilization of these programs are often inadequate. This study aimed to investigate community perceptions of school-based mass drug administration programs for these infections in Nigeria and to identify the barriers to their utilization and coverage.

**Methodology/Principal Findings:** The study used a qualitative research approach, involving focus group discussions and in-depth interviews with stakeholders involved in neglected tropical disease control programs in Ogun State, Nigeria. A semi-structured questionnaire was used to guide the exploration of ideas, and the data were analyzed using the QRS Nvivo 12 software package. The study found several barriers such as poor drug acceptability, accessibility, and effectiveness, low knowledge and awareness of the diseases and control interventions, inadequate community engagement and involvement, and weak health system and partner support to the utilization and coverage of control interventions for soil-transmitted helminths and schistosomiasis. The study also identified recommendations for addressing these barriers, including community sensitization and engagement, improving drug distribution and effectiveness, strengthening health system support, and enhancing partner collaboration and coordination.

**Conclusions/Significance:** The study revealed correct perceptions of transmission but some misconceptions about disease causation, transmission, and drug safety. Participants expressed a desire for better sensitization campaigns and more assurances of their safety. To improve mass drug administration programs, the study recommends strengthening health education messages and increasing the visibility of on-site medical personnel. The findings have implications for improving the effectiveness of these programs and reducing the burden of intestinal parasitic infections in the community. The study highlights the need for community engagement and education, health system support, and partner collaboration to ensure the successful implementation of mass drug administration programs.

**Author Summary:** This study explored the barriers to the utilization and coverage of control interventions for soil-transmitted helminths and schistosomiasis in Nigeria. Key informant interviews and focus group discussions were conducted with stakeholders involved in Neglected Tropical Disease school-based control programs, including community members, teachers, parents, and school-aged children. The study found that the main barriers to the utilization and coverage of control interventions for soil-transmitted helminths and schistosomiasis were poor drug acceptability, limited accessibility to drugs, and inadequate knowledge about the diseases and the control interventions. Additionally, the study found that the implementation of Neglected Tropical Disease control programs was inconsistent due to a lack of support from partners. Overall, our study provides important insights into the barriers to Neglected Tropical Disease school-based control programs and highlights the need for improved drug acceptability, accessibility, and knowledge about the diseases and control interventions. Our findings can inform the development of effective interventions to improve the utilization and coverage of control interventions.

## Introduction

Soil-transmitted helminths (STH) and schistosomiasis (SCH) are among 20 neglected tropical diseases (NTDs) identified by WHO and are responsible for approximately 150,000 deaths a year (1-3). Africa accounts for 40% of the global burden of NTDs (4). In several developing countries, NTDs are major public health problems including Nigeria. Nigeria has the largest burden of NTDs in sub-Saharan Africa, accounting for 25% of the total burden in the region (2, 5). In Nigeria, STH and SCH infections harm the overall health status and fitness of school-aged children (SAC) (6). STH and SCH are poverty-related diseases, and SAC is the group at the highest risk.

Over the last three decades, considerable attention and funding have been given to NTDs, with most recent efforts being targeted towards the attainment of commitments detailed in the WHO Ending the neglect to attain the Sustainable Development Goal that pursues the control and elimination of selected NTDs by 2030 (7, 8). STH and SCH are primarily controlled through mass drug administration (MDA) programs delivered to SAC via school-based delivery platforms to reduce the occurrence, severity, and long-term consequences of morbidity (9). While MDA is the primary strategy for the control and elimination of STH and SCH, the treatment has been inconsistent. There are concerns regarding the ability of these intervention achievements to be maintained in the face of programmatic, social, and political changes (2, 10). With the approaching year 2030, it is imperative to assess both the accomplishments and obstacles encountered in the implementation of MDA to ensure that the goal of equitable achievement of the ‘end game’ is attained in varied and rapidly evolving contexts.

A better understanding of the facilitators and barriers of integration within existing integrated programs can enhance public health integration efforts in Nigeria. The integration of NTD control programs has been identified as a highly promising approach toward achieving the London Declaration goals and has been recommended by the WHO for NTD-endemic countries to optimize program implementation (9). Since STH and SCH NTD control programs rely on MDA, it is crucial to comprehend the factors that impact the different stages of implementation, such as acceptability, coverage, and compliance. Moreover, identifying and assessing the factors that influence implementation outcomes is essential to ensure the successful implementation of NTD control programs. In the past, separate MDA programs were implemented for schistosomiasis and STH control in Ogun State. However, an integrated school-based MDA program for both diseases was introduced in 2016 (11). Despite significant progress in eradicating schistosomiasis and STH, there is a need to investigate the implementation challenges that hinder the uptake, utilization, and coverage of the integrated MDA programs at the community level. Understanding the community’s perceptions can provide valuable insights for program managers to improve current control strategies(12).

The primary aim of this study was to understand the community’s perceptions of the school-based MDA control programs for STH and SCH infections and how these perceptions could impact the control and elimination of these infections. Moreover, as the primary targets of the school-based MDA control programs, the SAC themselves possess knowledge, attitudes, and practices that must be understood and corrected to help eliminate these infections. Therefore, the study also includes exploring the community’s perceptions of SAC regarding the MDA control programs.

## Methods and materials

### Ethical statement

The study was conducted following the principles outlined in the Declaration of Helsinki, and all participant data was treated with strict confidentiality throughout the study. The study received ethical approval from the Institutional Review Board of the Nigerian Institute of Medical Research, Lagos, Nigeria (reference number IRB/21/004, approval date February 16, 2021), and social approval from the Department of Health Planning, Research, and Statistics, Ogun State Ministry of Health, Ogun State, Nigeria (reference number HPRS/351/388, approval date June 16, 2021). Verbal informed consent was obtained from all study participants after the objective and purpose of the study were explained to them. Informed assent forms were provided to children and minors, and informed parental consent was obtained for their parents or local guardians. For illiterate participants, impartial witnesses were enlisted to assist with the consenting process. Participants were included in the study only after they had individually signed the informed consent/assent form and received a paper copy of it.

### Study settings

Ogun State, Nigeria was selected as the study location due to its high prevalence of STH and SCH, and the inconsistent support of partner programs for NTD control. Four local government areas (LGAs) in Ogun State were purposively selected based on recommendations from the Department of Public Health, Ogun State Ministry of Health. The selected LGAs were Ikenne, Abeokuta North, Abeokuta South, and Obafemi Owode.

### Study Design and participants

This study employed a qualitative research approach to explore barriers to the utilization and coverage of control interventions in neglected tropical disease (NTD) control programs. The study focused on stakeholders involved in NTD control programs in Ogun State, Nigeria, where STH and SCH are endemic. Purposive sampling was used to select key informants and focus group participants based on their engagement in various positions in the health system and community, as well as their involvement in the management of NTDs. The study conducted in-depth Key Informant Interviews (KIIs) with community members such as the NTD focal persons. Additionally, the study conducted Focus Group Discussions (FGDs) with parents, teachers, town announcers, opinion leaders, enrolled school-aged children (ESAC), and non-enrolled school-aged children (NESAC). A semi-structured questionnaire was used as a tool during these sessions to guide the exploration of ideas. Thematic analysis was used to extract key themes and sub-themes from the data. This methodology enabled a comprehensive understanding of the challenges faced by NTD control programs in addressing STH and SCH and provided valuable insights into the barriers to achieving effective control interventions.

### Sample size determination

As a qualitative study, the authors did not perform sample size calculation; instead, they predetermined the number of Focus Group Discussions (FGDs) and Key Informant Interviews (KIIs) necessary to achieve ideal saturation. The authors planned to conduct 12 FGDs and 15 in-depth interviews to reach this goal.

### Data collection

To ensure the reliability of the data collected, the semi-structured and FGDs interview topics were developed and pretested (See Supplementary Materials 1-3). Audio tape recorders were used to capture all the information obtained during the KIIs and FGDs. The interviewers also made observations and took notes while recording to ensure accuracy. The interviews were conducted in both English (the national language of Nigeria) and Yoruba (the local language of the study participants) in various settings, including small halls, private rooms, and offices. The duration of each KII ranged from 20 to 40 minutes. FGDs were conducted in the participants’ respective LGAs.

### Data processing and analysis

Data from FGDs and KIIs were transcribed, coded, and analyzed thematically based on the emerging themes of barriers, drugs, knowledge, and recommendations. The recorded audio in the Yoruba language was transcribed into the English language. The transcription was checked independently by the team members for verification and accuracy with simultaneous audio playing. The study content was analyzed thematically, indexed, and coded inductively using the QRS Nvivo 12 software package (QRS International, Doncaster, Australia) for the content analysis of unstructured qualitative data. The initial open codes were sorted into sub-themes based on their similarity. These sub-themes were clustered and refined to form broad themes and debated within the research team. The nodes under drugs were categorized into three (acceptability, accessibility, and effectiveness) according to the participant’s responses. The knowledge nodes were categorized into awareness, causes, drugs, MDA, prevalence, mode of transmission, symptoms, and treatments. Data collection, analysis, and reporting followed the Standards for Reporting Qualitative Research (SPQR) guidelines.

To ensure consistency during protocol preparation, data collection, development of a coding system, interrater reliability, and data analysis, Guba and Lincoln’s criteria for determining rigor in qualitative research were used (13). Two interviews per group were coded by two authors (FT, DN), for which the degree of similarity was determined by calculating the interrater reliability using the QRS Nvivo 12 software package (QRS International, Doncaster, Australia). Cohen proposed that Kappa scores should be interpreted using the following: values ≤ 0 indicate lack of agreement, values 0.01 – 0.20 indicate no or little agreement, values 0.21 – 0.40 indicate fair agreement, values 0.41–0.60 indicate moderate agreement, values 0.61– 0.80 indicate substantial agreement, and values 0.81 – 1.00 indicate nearly perfect agreement. Kappa’s score for the thematic analysis was 1.00, indicating nearly perfect agreement.

### Data quality control

To ensure the quality of data, the KIIs and FGDs questions were well discussed by the team members before actual data collection. An intensive three-day training was given for data collectors on how to conduct in-depth interviews and FGDs. Supervision was done throughout the data collection process by the investigators.

## Results

The findings can be broadly split into four thematic areas, knowledge, barriers, drugs, and recommendations as shown in Table 1. Key components of MDA-integrated control programs such as the critical role of teachers, parents, health workers, and community members in engaging for MDA in terms of mobilization and sensitization were described by the study participants.

**Table 1.** Themes and Sub-themes.

| Themes | Sub-themes |
| --- | --- |
| **Knowledge** | 1. Perception of transmission<br>2. Perception of symptoms<br>3. Misconceptions about disease causation, transmission, and drug safety<br>4. Drug/Mass Drug Administration |
| **Drugs** | 1. Acceptability<br>2. Accessibility<br>3. Effectiveness |
| **Barriers** | 1. Low knowledge and awareness of the diseases and control interventions<br>2. Inadequate community engagement and involvement<br>3. Weak health system and partner support |
| **Recommendations** | 1. Community sensitization and engagement<br>2. Improving drug distribution and effectiveness<br>3. Strengthening health system support<br>4. Enhancing partner collaboration and coordination |

### Demographic Characteristics of Participants

A total of 144 community participants were involved in the study. A total of 12 FGDs were conducted amongst the community groups who have enrolled SAC and Non-enrolled SAC; teachers and parents; opinion leaders and town announcers. Table 2 below presents the composition of the KII and FGD and Table 3 the socio-demographic characteristics of the study participants in the KIIs and FGDs.

**Table 2.** Composition of Key Informant Interview and Focused Group Discussion.

| Participants | Female | Male | Total |
| --- | --- | --- | --- |
| Enrolled and non-enrolled school-aged children | 31 | 17 | 48 |
| Teachers/Parents | 31 | 17 | 48 |
| Community members | 26 | 22 | 48 |

**Table 3.** Socio-demographic characteristics of the study participants in the KIIs and FGDs.

| Description | Frequency<br>(N = 144) | Percentage<br>(%) |
| --- | --- | --- |
| **Gender** |  |  |
| Male | 56 | 39.0% |
| Female | 88 | 61.0% |
| **Age in years** |  |  |
| 6-9 | 8 | 5.60% |
| 10-14 | 30 | 20.8% |
| 15-19 | 10 | 6.90% |
| 20-24 | 9 | 6.30% |
| 25-29 | 8 | 5.60% |
| 30-34 | 25 | 17.40% |
| 35-39 | 20 | 13.9% |
| 40-44 | 18 | 12.5% |
| 45-49 | 12 | 8.30% |
| ≥ 50 | 4 | 2.80% |
| **Marital Status** |  |  |
| Single | 62 | 43.1% |
| Married | 77 | 53.5% |
| Divorced | 5 | 3.50% |
| **Religion** |  | 0.0 |
| Christianity | 79 | 54.9% |
| Islam | 60 | 41.7% |
| Missing | 5 | 3.50% |
| **Occupation** |  | 0.0 |
| Enrolled SAC | 24 | 16.7% |
| Non-enrolled SAC | 24 | 16.7% |
| Parents | 24 | 16.7% |
| School teachers | 24 | 16.7% |
| Opinion leaders | 24 | 16.7% |
| Town announcers | 24 | 16.7% |

### Theme 1. Knowledge

#### Sub-theme 1.1. Perception of transmission

Parents, teachers, ESAC, and NESAC were assessed on their knowledge of STH and SCH as well as the mode of transmission. Most of the parents, teachers, opinion leaders, and town announcers showed an acceptable knowledge of these diseases, and the mode of transmission was well highlighted by the teachers. The knowledge of STH and SCH and the mode of transmission was observed to be poor among the ESAC and NESAC.

A NESAC participant stated that schistosomiasis:

> *“It’s a disease that worries the stomach if one hasn’t eaten.”*

Another NESAC opined that,

> *“It’s something that worries the stomach if the drug isn’t being used properly”*

The most common response to the cause of STH infection was,

> *“do not pee in the river to prevent”*

Another NESAC said that,

> *“Not good to pee around, especially where the dog has peed. It will lead to bloody urine.”*

While another said the source of schistosomiasis infection was:

> *“contacted through water if an infected person has previously peed inside a river another human can contact it”*

An ESAC said that,

> *“worm causes malaria. It’s a bad disease that happens in Nigeria”*

Another ESAC said that,

> *“What I know about worm disease is that it does not allow our body to react the way it should. It also caused vomiting and dry body”*

When asked about the mode of transmission of these infections, an ESAC said that,

> *“urinating in the river to prevent it we should not be urinating in the river again.”*

Another ESAC said that

> *“It is bad for someone to be urinating everywhere because wherever a dog urinates and a person goes to urinate there such an individual can be infected by passing out bloody urine.”*

It can be said that most of the N/ESAC participants opined that the disease is caused by contact with dog urine.

> *“by urinating on the same spot, a dog urinates”*
>
> *“By urinating in the same spot, a dog urinates and bathing inside an infected river”*

While a few others attributed it to the consumption of sugary things

> *“if one eats many sugary things and eats dirty things”*
>
> *“Eating sugary things in the morning”*

#### Subtheme 1.2. Perception of symptoms

The majority of the community members interviewed had adequate knowledge of the symptoms of schistosomiasis and soil-transmitted helminths. An ESAC gave an account of her personal experience when she had the disease,

> “…*when I had worm disease, I had stomach pain and was vomiting; a worm also came out from my anus”*

A teacher elaborated on the symptoms of the disease.

> *“…according to what they said, it doesn’t normally show on the face but you can see it when the person urinates with a touch of blood. Also, a fat person infected will be getting lean. The children at times maybe he agile in class, but he will not feel agile anymore he will be dizzy, reluctant, will not feel happy the behavior will change generally.”*

Another teacher stated that the disease influences the student’s academic performance, she mentioned that the students infected with the disease show sign of low assimilation.

> *“…children were vomiting the worms out, stunted growth, poor assimilation, blood in the urine, loss of urine”*

A NESAC stated that

> *“… yes, there is, a particular drug given to us by the government which is being used every 3 months”*

#### Subtheme 1.3. Misconceptions about disease causation, transmission, and drug safety

A parent had a major misconception about the diseases and their symptoms

> *“I have seen two. He first is my child. He always complained of stomach upset after eating and he seemed to always want to throw up. The second is my neighbor’s child who always screamed while taking a pee”*

When asked about the symptoms exhibited by children who have contracted the diseases, ESACs had a popular idea in their mind that it causes stomach aches.

> *“When I am sick with worm, I have a stomach ache and feel vomiting”*
>
> *“One goes to defecate all the time when infected with a worm.”*
>
> *“When infected with the worm, you urinate and vomit most times and sleep a lot.”*
>
> *“It causes the stomach to be a big, thin body and big head.”*

Parents had a better opinion of the symptoms exhibited by children who contracted the diseases however some wrong ideas given include:

> *“blood in the urine”*
>
> *“severe stomach ache (sugary things)”*
>
> *“Discharge during urination”*
>
> *“blood in urine, do not pee in the river to prevent”*

Some first-hand experience by some non-enrolled children on the symptoms that they experience.

> *“What I know about worm disease is that it does not allow our body to react the way it should. It also caused vomiting and a dry body*.
>
> *“vomiting, being irritated, various tummy complaints, Stony saliva in the mouth”*
>
> *“Stony saliva in the mouth, nausea, vomiting, running stomach, weight loss”*
>
> *“When I had worm disease, I had stomach pain and was vomiting; a worm also came out from my anus. My dad took me to the hospital.”*

#### Theme 1.4. Mass Drug Administration

The MDA program was well accepted by the community. One respondent said:

> “*…Socialization meetings are being held every month, there’s good cooperation between us about the MDA.”*

An opinion leader also confirmed that the community members cooperate with them to make the MDA program effective. He stated that:

> *“*…*no direct communication, but there are representatives we get through. But there’s effective cooperation”*

Another respondent stated that:

> “*Through the records, it’s of high acceptability, and there has been positive support from people in the community.”*

A teacher who was involved in the program shared her experience which shows that there had been improvement in the MDA programs.

> “…*Eh! Well… it is effective, the reason I said that is that last year they give me the drug about 400 and something, but it is only 100 they used because they reject it but this time to God be the glory, I worked hard, talked to the parents on the benefits so now it is effective*.”

The reasons mentioned for poor mass drug administration first was that people heard that the drug killed a student who took the drug. An ESAC stated that,

> *“… a primary 4 pupil brought noodles. She slept after taking the drugs till closing time, and the next day we were told she died”*

When an opinion leader was asked about the incident during the FGD sessions, he said that it was a rumor which was later debunked.

> *“…though there were rumors and rejection, people were later educated on the drugs, and it’s always administered in the morning not far from breakfast.”*
>
> Another respondent, a parent also confirmed it and emphasized that the drug was given to them at no cost.
>
> *“ …government gives drugs which are not being sold just for worm alone and for schistosomiasis”*

Regarding the knowledge of the drug given to them, the majority of the ESAC and NESAC spoke about the color of the drug but only a few of them were able to mention the drug’s name. Although, the teachers and some parents were able to provide the names of the drugs.

A NESAC student stated that,

> *“The administrators ask about our ages and they measure our heights which then corresponds to the type of drug we will be given. Generally, these tablets are of different shapes: one is a little bit longer while the other is round in shape*”.

An ESAC stated that,

> “…*it’s white in color and long and the name of the drug is Albendazole”*

Furthermore, the ESAC and NESAC cited that before they can take the drug they are supposed to seek their parent’s consent. An ESAC said that,

> *“…our teachers instructed us to bring solid food to school. Mother was aware. Took ‘Eba’ to school. One minute after food, we were measured and drugs were given according to age and height. It was not easy to swallow.”*

Although few of the parents insisted that their child should not be given the drugs due to the side effects, a parent said:

> “…*there are some side effects but it’s being effective and successful, it’s a very powerful drug and if one didn’t eat a solid food it may cause dizziness or faint”*

According to an opinion leader, there are different drugs given for STH and SCH. He mentioned some of the drugs given.

> *“… Mectizan for eye worms, Albendazole for worms, mebendazole for schistosomiasis. When the disease is cleared people will be free to do what they want to do”*

Another opinion leader said,

> *“…before drugs, they should ensure they eat well because the drug is strong”*

Contrary to the treatment provided by the government, the respondents also mentioned the alternative treatment available in the community. A parent said,

> *“… people get treated, through herbal treatment (Yoruba herbs) and clinic care”*

Another parent explained further the alternative treatment provided by their traditional healers. She believed that although the treatment works there is a problem related to the usage of herbs as its dosage cannot be measured compared with clinical prescription. This view is depicted in a quote by a parent.

> *“The herbs, we have our traditional healers that they are using herbs for it*.
>
> *They cook and drink it without measurement and as they are drinking it, it will just come out through urine or feces.”*

Another parent also felt that,

> *“…although I always use the modern method because we have been advised to do so*.
>
> *Locally made drugs do not have dosage and that might even cause more harm to our children than good. I, therefore, believe that modern medication is the right medication for these diseases”*.

An opinion leader said,

> “*…I do not go for herbal concoction. In the hospital, there’s no significant cost, just card fees”*

### Theme 2: Drugs

#### Sub-theme 2.1. Drugs acceptability

Beliefs and perceptions about the drugs and their side effects were found to be critical in shaping acceptance. When asked about their opinion on drug acceptance in their community. The participants gave the following reasons.

> *“So the worm won’t worry them again, we will accept”*
>
> *“not to get infected with it again or come in contact again”*

A participant mentioned that he accepted it because a friend accepted it, and there were no side effects.

> *“……because she has used it before and didn’t worry her”*

However, some NESAC said they would not accept the drug stating some reasons which were mostly related to the side effects of the drugs. Some of the side effects highlighted were:

> *“fear of vomiting, bitter taste, and laziness”*
>
> *“Some people think the drug might make them vomit or stool”*
>
> *“Some set of people that received these medications complained of headache”*
>
> *“Some people who are already on medications refused to take these drugs and also, some felt taking the drugs will make them weak”*

From the responses of the NESAC, it was observed that most of them were skeptical about the drug and its side effects. Also, the ESAC shared similar sentiments on the side effects of the drug. For instance, a participant reported an adverse effect a brother had after using the drug.

> *“because it made my brother faint which made him come back home the next day, and she was warned by her people not to use it again”*

Acceptability was influenced by the decision of the mother. The following quotes from the N/ESAC demonstrate this,

> *“Last year she took it and fell ill, so her mother said No”*
>
> *“Because their mum asked them not to use it, that it will make them fall sick”*

FGDs among the ESAC and NESAC revealed rumors and parents’ opinions have a strong effect on drug acceptability.

Among some parents and teachers, it was observed that the drug has high acceptability as a parent stated:

> *“…*..*everyone in our community is using it. Even my kids”*

Another parent said

> *“yes everyone in my community is using it”*

And similarly, the opinion leaders thought that the drug is of high acceptability in their various communities as they all opined that

> *“Through the records, it’s of high acceptability, and there has been positive support from people in the community.”*

#### Sub-theme 2.2. Drug accessibility

The drugs are available and only administered by the health officers who summon parents to a meeting before administering them in the school. A teacher expressed,

> *“…*..*Yes, I believe they are health officers. They usually call for parents meeting before school distribution.”*

Another teacher said,

> *“…*..*right now the drug is available because we have been giving it to the children in school according to their age and height.”*

#### Sub-theme 2.3. Drug effectiveness

The opinion leaders gave their perceptions on the accessibility and effectiveness of the drugs administered, despite the identified barriers, they could attest to the efforts of the government as being effective to a level. An opinion leader expressed,

> *“Yes they are given and it’s being done through the house-to-house administration and also through mobilization to health centers”*

Another one stated with emphasis the personal effort embarked on to ensure that the drugs were effectively distributed in their location.

Similarly, another opinion leader reiterated this effort saying,

> *“Yes, we distribute ourselves into groups to increase coverage. Some go in the morning, some do afternoon in rounds.”*

However, the effectiveness of the drug was attested to by most of the opinion leaders. They mentioned the effectiveness of the drug as the reason why the drug is still being accepted for usage by some parents.

> *“There is a big effectiveness of the drug cause if there aren’t people won’t collect the drugs to use”*
>
> *“Yes, it is very effective, though it made some kids dizzy it’s very effective.”*

A parent acknowledged there were side effects though the drug is effective,

> *“though there are some side effects it’s effective and successful, it’s a very powerful drug and if one didn’t eat solid food it may cause dizziness or fainting. And in case of that, they are told to use glucose”*

### Theme 3: Barrier

#### Theme 3.1. Low knowledge and awareness of the diseases and control interventions

Perceived barriers were assessed among the study participants to the acceptability of the drugs. Responses from the ESAC and NESAC were consistent. ESAC noted that,

> *“…people decline because of parents’ instructions because their mum asked them not to use it that it could make them sick.”*
>
> *“…. the fear of vomiting and laziness, some children throw the drug away because it is bitter”*
>
> *“…the smell of the drug, it is not nice”*

A NESAC opined that,

> *“…. some parents warned their children not to receive these medications, saying the medications are bad”*
>
> *“…*.. *the smell of drugs is not nice, I fear vomiting, bitter taste, and laziness”*

In the FGDs among the parents, it was mentioned that,

> *“…the parents are ignorant; they don’t know what the drug is about.”*

Others stated the side effects of the drugs as barriers to the acceptability of the drug. Interestingly, a parent emphasized an important factor that affects the acceptability of the drug to her and some others.

> *“Appearance shows the manner. Sometimes the medical staff appears moody and all sorts to the parents and people. This will discourage the masses from accepting what is being brought to them”*

#### Theme 3.2. Inadequate community engagement and involvement

The perspective of opinion leaders and town announcers were sought after being the gatekeeper of the communities. They gave several reasons that could serve as a hindrance to the acceptability of the drug which would help assist the providers to strategize in the distribution of the drug.

Transportation of the drug was highlighted as one of the barriers experienced. Another potential barrier identified was the late announcement/sensitization of the MDA roll-out.

> *“…. Time allowed for campaign and education is not enough.”*

Another participant noted that incentives given to drug administrators are important:

> *“The small incentive is discouraging. When there was no payment, we didn’t go around, but the turnout was poor. People said we didn’t give them food to take medicine. But now we do house-to-house. The money is small no doubt, but if it is paid on time it will be encouraging.”*

A participant noted that rumors about the drug were a deterrent to the acceptability of the drug stating that:

> *“These things give a draw-back to this program because some of these rumors are not true and didn’t happen”*

#### Theme 3.3. Weak health system and partner support

Some opinion leaders opined that the drug administered from a private health facility will give it more credibility. Also, it was the purchasing power of the parents when they want to buy outside of the period of MDA by the state government. Someone mentioned that some religious leaders preach against the usage of drugs. This is a vital issue to be explored by the MDA providers.

### Theme 4

#### Themes 4.1. Community sensitization and engagement

Several recommendations were given to improve the MDA program. NESAC mentioned the importance of financial incentives to people for using the drug.

> *“Bribing people will ensure the usage.”*
>
> *“give kids incentives”*
>
> *“Distribution should be followed by incentives, which can be distributed through schools as kids fear the teachers.”*

Similarly, ESAC also suggested giving incentives.

> *“Sweets and candies should be shared with the children while distributing the drugs to the children, they should be cajoled into taking it by promising the nice things e.g. biscuits, sweets and telling them the drug is sweet”*
>
> *“By buying things for the children; biscuits sweets… even the reluctant children will accept the drug afterward”*

Some others opined that,

> *“Awareness through radio programs and media for people to know the efficiency and effectiveness of the drug”*
>
> *“by educating them about the drug for them to know about the effectiveness”*

#### Themes 4.2. Improving drug distribution and effectiveness

Interestingly, another NESAC suggested that the drug be made more palatable to use in combination with effectiveness and parents influencing their kids to use.

> *“…. if the drug is sweet if it works well in the body if parents insist they use it.”*

Some recommended educating parents and even the kids on the benefit of taking the drug.

> *“Inform the child and parents about the effectiveness of the drug”*
>
> *“When you educate and sensitize them and also give them gifts”*

Parents’ and teachers’ recommendations were captured under the major theme of sensitization. Parents encouraged the government to increase the sensitization of the public on the drug and the program.

> *“Government should continue to do more awareness to people”*
>
> *“Awareness creation and educating them more, on media about the drug”*
>
> *“Continue doing the publicity because most parents are ignorant that is the problem.”*

#### Themes 4.3. Strengthening health system support

Some proactive measures were recommended by some parents:

> *“Appearance shows the manner. Sometimes the medical staff appears moody and all sorts to the parents and people. This will discourage the masses from accepting what is being brought to them”*
>
> *“The medical staff should try to show love and joy while passing information across and try to explain the benefits very well so people can understand”*

Other parents recommended food during the drug distribution.

> *“Availability of food for them to use the drug”*
>
> *“….If they can extend the time and continuity of the drug and ensure availability of food during the drug administration”*
>
> *“To ensure there’s the availability of food because the drug is strong”*

Opinion leaders gave similar recommendations in the same vein that awareness should be done so rigorously.

> *“megaphone should be given to creating awareness”*
>
> *“Proper awareness through media to encourage people.”*
>
> *“Education of the drug to people”*
>
> *“They should make awareness and the drug should be made available and brought to every one school and house”*

Some opinion leaders opined that the welfare of the frontline workers is taken more seriously.

> *“The health care workers should be compensated and paid well for easy work and effective awareness”*
>
> *“it’s to ensure the health workers are paid and proper awareness should be made.”*
>
> *“I will advise they pay health workers and take all things reported with all seriousness”*
>
> *“mobilizers need money, transportation too. Everything needs to be funded, even the megaphones we use need a battery, so you can see money is key”*
>
> *“Government shouldn’t relent and ensure funds are paid and do more of awareness creation”*

Some opinion leaders even recommended that MDA administrators take the drug in the presence of the student to encourage them to use it.

> *“The drug administrators should use the medicines first”*

Another opinion leader recommended that there should be a repeat of the exercise for students who missed the day of administration.

> *“Mectizan should be twice a year because some are upset when they miss the drug distribution due to travel”*
>
> *“The interval is too long, adequate education is required before administration”*

## Discussion

Mass Drug Administration for vulnerable populations has been the best approach for the prevention of STH and SCH globally (14, 15). Our findings from key stakeholders and school-aged children highlighted four main areas: knowledge, drug acceptability, accessibility, barriers, and recommendations. The participants were drawn from different interest groups whose socio-demographic characteristics demonstrated the appropriateness of each category. It was important to find out if the children, the recipients of the drugs were aware of the knowledge of the drugs and the process of MDA. It was difficult for the ESAC and NESAC to differentiate between STH and SCH infection and their mode of transmission. The inadequacy of knowledge among the SAC has been substantiated by evidence from several studies in Africa for instance studies done in Malawi, Kenya, Uganda, and Tanzania (16-20). Poor environmental and sanitary conditions are linked to SCH infections (21) while STH is transmitted through contaminated soil (22, 23). Our findings revealed that most participants associated these diseases with drinking dirty contaminated water (17). There were several misconceptions like transmission through urinating where dogs have urinated and bathed inside infected rivers. These findings agree with an Ethiopian study on low awareness and common misconceptions about schistosomiasis (24). Limited knowledge has been highlighted as a factor promoting the transmission of STH and SCH infection (25).

The majority of the participants demonstrated adequate knowledge of the symptoms of STH and SCH infection. They pointed out bloody stools and urine, vomiting, fever, headaches, abdominal aches, and generalized body aches.

It can be assumed that with a good knowledge of the drugs and the potential benefits, the acceptance would be better and the attitude towards it would improve. In regards to the treatment of SCH and STH, the majority of the respondents were aware of the importance of the treatment and they affirm that the government provides drugs for them. The provision of drugs by the government is an advantage to these communities as there is literature contrasting this with reports of drug shortages as challenges being encountered (26, 27). The children only recalled the color of the drugs in contrast to the adult respondents who were able to state that there were different drugs given for STH and SCH.

Both STH and SCH were acknowledged as a burden among the community members especially due to the knowledge deficit. This is echoed in several studies on the knowledge, attitudes, and practices of SCH and STH (17, 18, 26, 28-30).

It was evident that people were involved in health programs in one way or another. The study revealed that the people in the community understand the implication of schistosomiasis and soil-transmitted diseases which makes them cooperate more and make the MDA program effective thus underscoring the impact of MDA in the control of SCH and STH (23, 31).

The acceptability of the administered drug during the MDA program was assessed among the ESAC and NESAC by asking if the children would accept the drugs or not. It was noted that the majority of the NESAC accepted the drugs because of the benefit of the drug. Some others did not accept the drug based on the color, bitterness, or bad taste and others mentioned side effects like headaches. These findings are in line with Kimani et.al. on the acceptability of praziquantel in the treatment of *Schistosoma haematobium* in preschool children (32). The family has a great influence on the acceptability of MDA(33), our findings revealed that the decision to accept the MDA was based on the influence of the mothers. On the accessibility, the children could not tell categorically how it could be accessed, however, the parents and the teachers expressed that the drugs were accessible in their various communities and were only administered by the health officers who summoned the parents to a meeting in school before the MDA exercise.

The parents and teachers voiced that fears about the drugs and acceptability of the drug are a hindrance to the acceptance and uptake of the MDA program among them even as stakeholders to the children. The majority of parents and teachers noted that there is a need for further enlightenment on the program and the effects of the drug especially. Further, our findings revealed drug side effects, parental instructions against usage, and unpalatable drug characteristics as barriers. Similar barriers have been identified in other studies(26, 31, 34).

Some recommendations were fronted by the study participants; categorized as improving the welfare package of the frontline health workers on the program, revisiting of distribution strategy, increasing awareness levels, and giving incentives to children. To increase effectiveness, there is a need for clear, concise, and consistent sensitization to the communities (17, 28, 31).

In this community perspective study, we found that participants generally had correct perceptions of STH and SCH transmission. However, some misperceptions persisted regarding disease causation and transmission, as well as fear and distrust surrounding the deworming drugs used in mass drug administration (MDA). Specifically, participants were concerned about how to handle side effects and adverse effects, the capabilities of MDA providers to address them, and perceived inferiority to commercially available anthelmintic drugs. Despite these challenges, participants acknowledged a knowledge gap and expressed a desire for better sensitization campaigns, information dissemination, and assurances of their safety. To address these issues, we recommend strengthening health education messages and increasing the visibility or availability of on-site medical personnel. By understanding how parents and children perceive MDA activities, policymakers and program implementors can work to eliminate barriers to compliance and achieve the target coverage rate for administering chemo-preventive drugs for intestinal parasites. Overall, our findings have important implications for improving MDA programs and increasing their effectiveness in reducing the burden of intestinal parasitic infections in this community.

The study had some limitations. Firstly, it was conducted in communities in Ogun State, Nigeria and therefore the findings may not be generalizable to other populations or communities with different socioeconomic or cultural backgrounds. Secondly, participants may have provided socially desirable responses, leading to potential social desirability bias. Thirdly, the study relied on self-reported data which may have been subject to recall bias or misreporting. Fourthly, the study only collected cross-sectional data at a single point in time, and longitudinal data would have allowed for a better understanding of changes in community perceptions and behaviors over time. Lastly, language barriers may have affected the quality and accuracy of the data collected, as the study was conducted in a language that may not have been familiar or comfortable for all participants.

## Data Availability

Yes - all data are fully available without restriction

## Acknowledgment

This publication was supported by a grant from the African Research Network for Neglected Tropical Diseases (ARNTD) through United States Agency for International Development (USAID), UK aid from the British people, and the Coalition for Operational Research on Neglected Tropical Diseases (COR-NTD). Its content is solely the responsibility of the authors and does not necessarily represent the views of ARNTD, COR-NTD, UK aid, or USAID.

## Supporting information

**S1 File. Key Informant Interview Guide**

**S2 File. Focus Group Discussion Guide for Parents, Teachers, Opinion leaders, and Town announcers**.

**S3 File. Focus Group Discussion Interview Guide for School-aged Children**.

## Notes

### Competing Interest Statement

The authors have declared no competing interest.

### Author Declarations

Institutional Review Board of the Nigerian Institute of Medical Research, Lagos, Nigeria (reference number IRB/21/004, approval date February 16, 2021), and social approval from the Department of Health Planning, Research, and Statistics, Ogun State Ministry of Health, Ogun State, Nigeria (reference number HPRS/351/388, approval date June 16, 2021).

